# Effects of amyloid and *APOE4* on medial temporal lobe subregions in cognitively unimpaired elderly

**DOI:** 10.1101/2022.01.20.22269607

**Authors:** Robin de Flores, Solène Demeilliez-Servouin, Elizabeth Kuhn, Léa Chauveau, Brigitte Landeau, Nicolas Delcroix, Julie Gonneaud, Gaël Chételat

## Abstract

Medial temporal lobe (MTL) sub-structures are differentially affected in early Alzheimer’s disease (AD), with a specific involvement of the entorhinal cortex (ERC), the perirhinal cortex (PRC) and CA1. However, the impact of amyloid (Aβ) pathology and APOE ε4 on MTL subregional atrophy remains relatively unknown. Our aim was to uncover these effects to further our understanding of the mechanisms underlying MTL atrophy in a population at-risk for AD.

We used baseline data from 130 unimpaired older adults (mean age: 68.9 ± 3.8 years) from the Age-Well randomized controlled trial for whom high-resolution structural MRI (T2-weighted; 0.4×0.4×2.5mm3), amyloid-PET (Florbetapir) and APOE genotype were available. Participants were dichotomized into amyloid positive (Aβ+, n=27) and negative (Aβ-, n=103), and APOE ε4 carrier (ε4+, n=35) and non-carriers (ε4-, n=95). Hippocampal subfield (CA1, CA2, CA3, dentate gyrus [DG], subiculum [SUB]) and extra-hippocampal region (ERC, Brodmann area [BA] 35 and 36, and parahippocampal cortex [PHC]) volumes were estimated using ASHS and normalized by total intracranial volume. For each subregion, group comparisons were performed (Aβ+ vs Aβ- and ε4+ vs ε4-) using ANCOVAs, including age, sex and education as covariates. Interactions with age (i.e., Aβ status * age and APOE ε4 status * age) were also investigated for each subregion.

No significant differences were observed between Aβ+ and Aβ-, nor between ε4+ and ε4-. However, a significant Aβ status * age interaction were observed for CA1 (p<0.05), where volumes were negatively associated with age in the Aβ+ group only. In addition, significant APOE ε4 status * age interactions were found for CA1, SUB, ERC, DG and the whole hippocampus (p<0.05), where volumes were negatively associated with age in the ε4+ group only.

Overall, our analyses showed that both Aβ and APOE ε4 status interact with age on CA1, which is known to be specifically atrophied in early AD. In addition, APOE ε4 status mediated the effects of age on other subregions (SUB, ERC, DG), suggesting a more important contribution of APOE ε4 than amyloid to MTL atrophy in cognitively unimpaired population. These results are particularly important to develop MRI-based biomarkers to detect early AD and further our understanding of the mechanisms underlying MTL atrophy.

## 1. Introduction

The medial temporal lobe (MTL) is one of the most studied brain region because of its involvement in a myriad of cognitive processes and its sensitivity to aging, neurological and psychiatric diseases, including Alzheimer’s disease (AD) (Squire et al., 2004; Geuze et al., 2005; Moscovitch et al., 2006; Small et al., 2011; Eichenbaum, 2017; Hodgetts et al., 2017). In the context of AD, histological studies have reported that the MTL is the first region to be targeted by neurofibrillary tangles (NFT), although MTL subregions are not affected homogeneously (Braak and Braak, 1991; Delacourte et al., 1999). NFTs first target the perirhinal and entorhinal cortices (PRC and ERC, respectively) and then target the hippocampal subfield CA1, then the subiculum (SUB), CA2, CA3 and the dentate gyrus (DG) (Braak and Braak, 1991, 1995; Lace et al., 2009). Importantly, magnetic resonance imaging (MRI) studies have consistently reported a similar pattern of atrophy, with an early involvement of ERC, PRC and CA1 in early AD stages (ie in patients with mild cognitive impairment (MCI)) while all MTL subregions appeared as affected at the AD-dementia stage (de Flores et al., 2015a; Gertje et al., 2016; Pini et al., 2016; Wolk et al., 2017; Adler et al., 2018; Hata et al., 2019; Xie et al., 2019). Thus, MTL atrophy is commonly used as a biomarker to monitor in-vivo AD-related neurodegeneration and provides important pathologic staging information (Jack et al., 2018). However, results are limited and less consistent in preclinical AD (ie amyloid-positive (Aβ+) cognitively unimpaired (CU)), with studies reporting atrophy in SUB/preSUB (Hsu et al., 2015; Parker et al., 2019), Brodmann area 35 (BA35 - a subregion of the perirhinal cortex (PRC)) (Wolk et al., 2017) or no atrophy (Xie et al., 2019) in comparison to Aβ-CU. The same conclusion applies to the effects of APOE polymorphism, the APOE ε4 allele being the largest genetic risk factor for sporadic AD, with limited and inconsistent observations (Vilor-Tejedor et al., 2021). For example, studies reported no effects of APOE ε4 on hippocampal subfields volume (Voineskos et al., 2015) while others reported atrophy of CA3/DG (Mueller et al., 2008) or CA1-SRLM (Kerchner et al., 2014). Since MTL subregional atrophy could be useful in the early diagnosis and monitoring of AD, it is particularly important to better characterize the deleterious effects of amyloid and APOE on these structures. Our aim was to uncover these effects to further our understanding of the mechanisms underlying MTL atrophy in the context of normal aging and AD. To that end, dedicated high resolution T2-weighted scans acquired in cognitively unimpaired elderly were used together with a tailored automatic segmentation algorithm.

## 2. Material and methods

### 2.1. Participants

One hundred and thirty cognitively unimpaired older adults from the baseline visit of the Age-Well randomized clinical trial of the Medit-Ageing European Project were included (Poisnel et al., 2018). Participants were recruited from the general population, older than 65 years, native French speakers, retired for at least 1 year, educated for at least 7 years, and able to perform within the normal range on standardized cognitive tests. They did not show evidence of a major neurological or psychiatric disorder (including alcohol or drug abuse), history of cerebrovascular disease, presence of a chronic disease or acute unstable illness, and current or recent medication usage that may interfere with cognitive functioning. All participants gave their written informed consent prior to the examinations, and the Age-Well randomized clinical trial was approved by the ethics committee (Comité de Protection des Personnes Nord-Ouest III, Caen, France; trial registration number: EudraCT: 2016-002441-36; IDRCB: 2016-A01767-44; ClinicalTrials.gov Identifier: NCT02977819). Each participant underwent a ^18^F florbetapir PET scan that reflect amyloid burden. A global SUVr measure was used to dichotomize the population into amyloid positive (Aβ+, n=27) and negative (Aβ-, n=105) based on a threshold defined in a group of 45 healthy young individuals younger than 40 years. In addition, APOE genotype was assessed and the cohort was also dichotomized into APOE ε4 carrier (ε4+, n=36) and non-carriers (ε4-, n=96). Demographics of the cohort are reported in Table 1.

**Table 1:** Demographics of the population.

| | Full cohort | $\text{A}\beta^-$ | $\text{A}\beta^+$ | $\epsilon 4^-$ | $\epsilon 4^+$ |
| --- | --- | --- | --- | --- | --- |
| N | 130 | 103 | 27 | 95 | 35 |
| Age (years) | 68.9 (3.8) | 68.5 (3.4) | 70.2 (4.7) | 69.0 (3.7) | 68.4 (4.0) |
| Gender (F/M) | 80/50 | 63/40 | 17/10 | 63/32 | 17/18 |
| Education (years) | 13.2 (3.1) | 13.3 (3.0) | 12.8 (3.4) | 13.2 (3.0) | 13.3 (3.3) |

### 2.2. MRI Data Acquisition and Handling

Each subject underwent an MR scan at the CYCERON center (Caen, France) using a 3T Philips (Eindhoven, The Netherlands) scanner. First, T1-weighted structural image were acquired (Repetition Time (TR) = 6.8 ms; Echo Time (TE) = 3.1 ms; flip angle = 6°; 180 slices; slice thickness = 1 mm; Field of View (FoV) = 256 × 256 mm2; matrix = 256 × 256; in-plane resolution = 1 × 1 mm2; acquisition time = 5min16). In addition, two high resolution T2-weighted structural images were acquired perpendicularly to the long axis of the hippocampus (TR = 5,310 ms; TE = 110 ms; flip angle = 90°; 23 slices; slice thickness = 2.5 mm; FOV = 140 × 111 mm2; matrix = 352 × 352; in-plane resolution = 0.398 × 0.398 mm2, acquisition time = 3min43). Then, both images were corregistrated and averaged using SPM12.

Hippocampal subfield (CA1, CA2, CA3, DG, SUB) and extra-hippocampal region (ERC, BA35, BA36, and parahippocampal cortex [PHC]) volumes were estimated automatically on the high-resolution T2-weighted images using the Automated Segmentation for Hippocampal Subfields software (Yushkevich et al., 2015). The volume of the hippocampus was estimated as the sum of the five hippocampal subfields. All volumetric measures were normalized to the total intracranial volume (TIV) to compensate for interindividual variability in head size.

### 2.3. Statistical Analyses

For each MTL subregion, group comparisons were performed (Aβ+ vs Aβ- and ε4+ vs ε4) using ANCOVAs, including age, sex and education as covariates. Interactions with age (i.e., Aβ status * age and APOE ε4 status * age) were also investigated for each subregion. Post-hoc analyses were performed in case of significant interaction (p < 0.05). Lastly, both the amyloid and APOE status were entered in the same model (including age, sex and education as covariates) and all analyses were repeated. This allowed us to assess whether specific effects of amyloid or APOE existed while controlling for the other one.

## 3. Results

No significant group differences were found between Aβ+ vs Aβ-nor between ε4+ vs ε4-(see Table 2). However, a significant interaction was found between age and amyloid status on CA1 (F=4.47, p=0.04). More precisely, CA1 was negatively associated with age in Aβ+ individuals (r=-0.48, p=0.01) but not in Aβ-individuals (r=-0.05, p=0.58) (see Figure 1).

**Table 2:** Effects of amyloid and APOE4 status on MTL subregions.

**a) Amyloid Status**
| | Volume<br>(TIV-normalized) | | Group comparison<br>(Aβ- vs Aβ+) | | Interaction<br>(Aβ status * age) | |
| --- | --- | --- | --- | --- | --- | --- |
| | Aβ- | Aβ+ | F | P | F | P |
| CA1 | 0.96 (0.10) | 0.93 (0.14) | 0.59 | 0.44 | **4.47** | **0.04** |
| CA2 | 0.02 (0.003) | 0.02 (0.004) | 1.70 | 0.19 | 0.58 | 0.45 |
| CA3 | 0.05 (0.01) | 0.06 (0.01) | 4.10 | 0.05 | 0.05 | 0.82 |
| DG | 0.42 (0.05) | 0.40 (0.07) | 0.54 | 0.46 | 1.95 | 0.17 |
| SUB | 0.32 (0.04) | 0.32 (0.04) | 0.55 | 0.46 | 1.20 | 0.28 |
| ERC | 0.41 (0.04) | 0.40 (0.07) | 0.63 | 0.43 | 3.56 | 0.06 |
| BA35 | 0.36 (0.06) | 0.35 (0.04) | 1.22 | 0.27 | 2.03 | 0.16 |
| BA36 | 1.24 (0.18) | 1.25 (0.19) | 0.39 | 0.53 | 1.47 | 0.23 |
| PHC | 0.67 (0.09) | 0.66 (0.09) | 0.05 | 0.82 | 3.68 | 0.06 |
| Whole hippocampus | 1.77 (0.18) | 1.73 (0.24) | 0.47 | 0.49 | 3.54 | 0.06 |

**b) APOE Status**
| | Volume<br>(TIV-normalized) | | Group comparison<br>(ε4- vs ε4+) | | Interaction<br>(APOE status * age) | |
| --- | --- | --- | --- | --- | --- | --- |
| | ε4- | ε4+ | F | P | F | P |
| CA1 | 0.96 (0.10) | 0.93 (0.13) | 1.06 | 0.30 | **4.31** | **0.04** |
| CA2 | 0.02 (0.003) | 0.02 (0.004) | 0.39 | 0.53 | 0.01 | 0.92 |
| CA3 | 0.05 (0.01) | 0.05 (0.01) | 0.91 | 0.34 | 0.47 | 0.50 |
| DG | 0.41 (0.05) | 0.41 (0.07) | 0.18 | 0.68 | 3.78 | **0.05** |
| SUB | 0.32 (0.04) | 0.32 (0.04) | 0.14 | 0.71 | **5.21** | **0.02** |
| ERC | 0.41 (0.04) | 0.40 (0.06) | 0.09 | 0.76 | **4.56** | **0.03** |
| BA35 | 0.37 (0.06) | 0.34 (0.06) | 3.82 | 0.05 | 0.46 | 0.50 |
| BA36 | 1.24 (0.16) | 1.26 (0.22) | 0.03 | 0.87 | 2.61 | 0.11 |
| PHC | 0.67 (0.08) | 0.66 (0.10) | 0.06 | 0.81 | 2.66 | 0.11 |
| Whole hippocampus | 1.77 (0.17) | 1.73 (0.24) | 0.76 | 0.39 | **4.97** | **0.03** |

**Figure 1:**
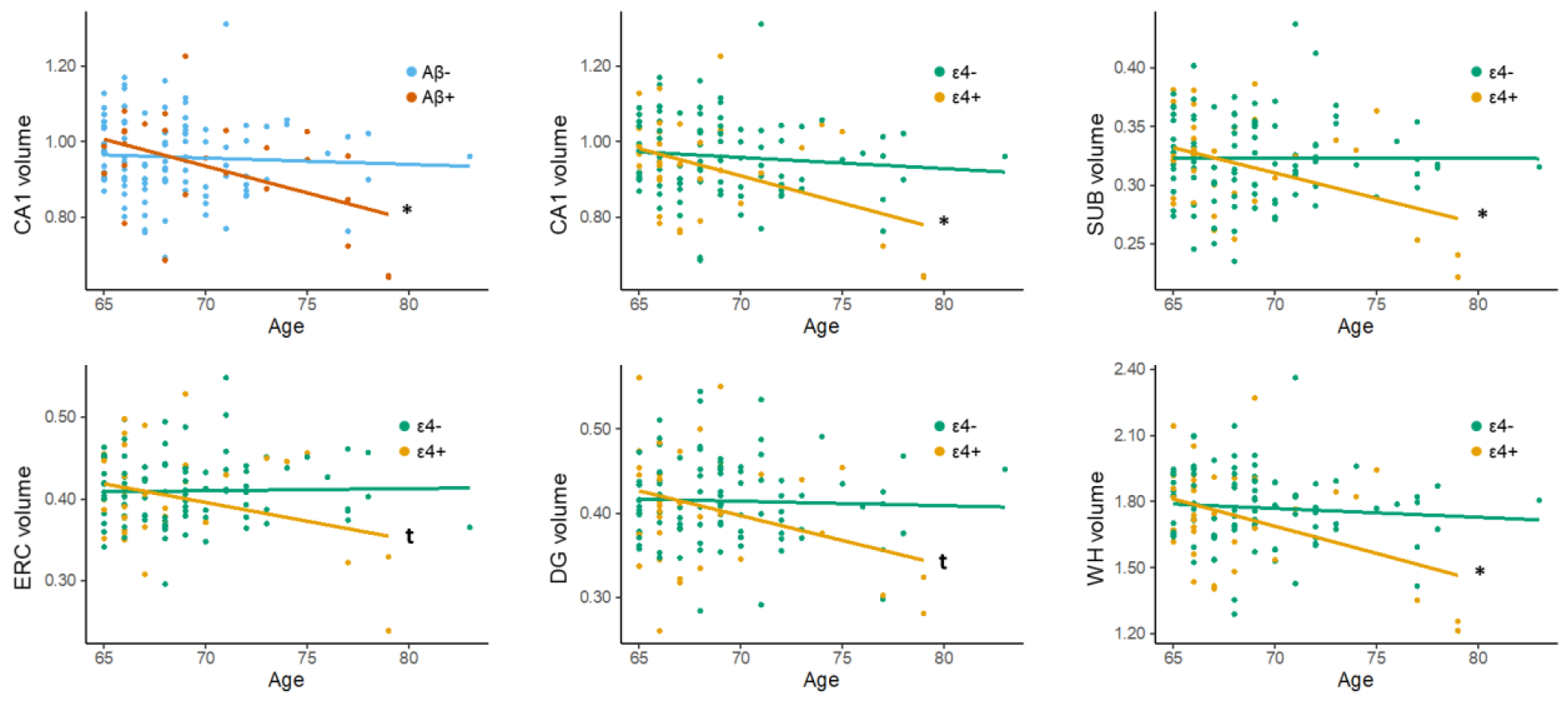
Significant interactions between Aβ status and age as well as between ε4 status and age on medial temporal lobe structures. All volumes were normalized by the total intracranial volume. SUB: subiculum; ERC: entorhinal cortex; DG: dentate gyrus; WH: whole hippocampus

In addition, significant interactions were found between age and APOE status on CA1 (F=4.31, p=0.04), SUB (F=5.21, p=0.02), ERC (F=4.56, p=0.03), DG (F=3.78, p=0.05) and the whole hippocampus (F=4.97, p=0.03). Post-hoc analyses showed that the volumes of CA1, SUB and the whole hippocampus were significantly associated with age in ε4+ subjects (r=-0.43, p=0.01 ; r=-0.41, p=0.01 ; r = -0.41, p=0.01, respectively), while trends were found for ERC and DG (r=-0.31, p=0.07 ; r=-0.33, p=0.06, respectively). In contrast, none of these regions were associated with age in ε4-individuals (r=-0.11, p=0.31 ; r=-0.01, p=0.97 ; r = -0.09, p=0.40 ; r=0.02, p=0.82 ; r=-0.04, 0.70, respectively) (see Figure 1).

Results were overall unchanged when both the amyloid and APOE status were entered in the same model (see supplementary table 1).

## 4. Discussion

In the present study, we used high resolution T2-weighted images together with a tailored automatic segmentation algorithm to investigate the effects of Amyloid and APOE4 status on MTL structures in cognitively unimpaired elderly. While we did not find significant differences in volume between Aβ+ and Aβ-nor between ε4+ and ε4-, our analyses revealed significant interactions between Aβ status and age as well as between APOE ε4 status and age on several MTL subregions.

Characterizing the contribution of amyloid to atrophy in cognitively unimpaired population is particularly important to design biomarkers that are sensitive and specific to early (preclinical) AD, especially since the disease tends to be defined biologically (Jack et al., 2018). Results from the literature are very heterogenous, with cross-sectional studies reporting atrophy of the hippocampus (Dickerson et al., 2009; Storandt et al., 2009; Whitwell et al., 2013; Nosheny et al., 2019) or BA35 (Wolk et al., 2017) in Aβ+ subjects as compared to their Aβ-counterparts, while other groups failed to report such differences (Bourgeat et al., 2010; Mattsson et al., 2014; Oh et al., 2014; Yushkevich et al., 2015; Chauveau et al., 2021). Regarding hippocampal subfields, two studies reported atrophy in SUB/preSUB (Hsu et al., 2015; Parker et al., 2019). However, the method used in these papers, which consisted in applying FreeSurfer to MRI scans with approximately 1 × 1 × 1 mm^3^ resolution, raised concerns (de Flores et al., 2015b; Wisse et al., 2021). Interestingly, Mueller et al. (2018) tested several image types and algorithms to detect amyloid associated subfield atrophy in cognitively normal subjects from ADNI. Using high resolution T2 hippocampal images, the authors found volume reduction in PHC and ERC with ASHS, and in SUB with FreeSurfer 6. Using T1 images, they did not find atrophy with FreeSurfer 5.1 but showed atrophy in CA1 using shape analysis. These inconsistencies certainly reflect the variability in the anatomical definition of hippocampal subfields across techniques but also suggest the inexistence of a reliable neuroanatomical signature of preclinical AD, as expressed by Whitwell et al. (2013).

Although we did not find direct group differences between Aβ+ and Aβ-individuals, our analysis showed a significant interaction between Aβ status and age on CA1, with this region being negatively associated with age in Aβ+ but not in Aβ-. Interestingly, this region has been extensively described as atrophied in early AD (de Flores et al., 2015a). To our knowledge, no other study have investigated the direct interactions between age and amyloid on hippocampal subfields. Interestingly, Wisse et al. (2022) recently showed that CSF p-tau levels partially mediated age effects on hippocampal atrophy rates. In addition, several studies using tau PET tracers showed higher MTL uptake in Aβ+ subjects as compared to Aβ- (Maass et al., 2017; Jagust, 2018; Pereira et al., 2020). Thus, the association between age and the volume of CA1 in Aβ+ participants could reflect the presence of tau pathology, especially since CA1 is the first hippocampal region to be targeted by tau in AD (Braak and Braak, 1991, 1995; Lace et al., 2009).

In addition to evaluating the contribution of amyloid to MTL atrophy, a second aim was to investigate the effects of APOE genotype on MTL structures. We did not find any significant differences between APOE ε4 carriers and non-carriers individuals. APOE ε4 is the largest genetic risk factor for sporadic AD and its effect on brain atrophy has been largely studied, with discrepant observations showing atrophy in AD-sensitive regions in some studies but contradicting results as well (Fouquet et al., 2014; Tzioras et al., 2019). For example, studies found significant differences in hippocampal volume between ε4 carriers and non-carriers (Tohgi et al., 1997; Lind et al., 2006; Wishart et al., 2006; Honea et al., 2009; Cacciaglia et al., 2018; Veldsman et al., 2021), while others did not (Reiman et al., 1998; Trivedi et al., 2006; Khan et al., 2017; Dong et al., 2019; Van Etten et al., 2021). Results are also inconsistent when specifically investigating hippocampal subfields atrophy. Indeed, studies reported volume reductions of CA3/DG (Mueller et al., 2008), CA1-SRLM (Kerchner et al., 2014), the molecular layer (Dounavi et al., 2020), SUB (Donix et al., 2010) in APOE ε4 carriers whereas others failed to report such differences (Burggren et al., 2008; Reiter et al., 2017). Interestingly, while they did not find differences in volume, Burggren et al. (2008) showed that cortical thickness was reduced in SUB and ERC among APOE ε4 carriers when compared to non-carriers. This observation suggests a greater sensitivity of thickness measures over volumetry to detect the effects of APOE, which has also been argued more recently by Dong et al. (2019).

While we did not find significant differences between APOE ε4 carriers and non-carriers, our analysis showed significant interactions between APOE ε4 status and age on CA1, SUB, ERC, DG and the whole hippocampus, with these structures being negatively associated with age in carriers only. Such interactions were previously described, especially in CA1 (Martí-Juan et al., 2021) or in all FreeSurfer 6 subfield ROIs (Veldsman et al., 2021). In a previous study, our group failed at reporting interactions between APOE ε4 status and age on MTL structures (Chauveau et al., 2021). However, it is important to note that this work was performed using standard T1 isotropic 1 mm^3^ images, where subfields were not investigated. Recently, several studies showed higher MTL tau-PET uptake in APOE ε4 carriers in comparison to non-carriers (Ramanan et al., 2019; Baek et al., 2020; Pereira et al., 2020; Therriault et al., 2020; La Joie et al., 2021; Salvadó et al., 2021). Note that while the APOE ε4 allele is thought to promote tau pathology, the mechanism underlying these effects may be indirect, mediated by APOE effects on microglia (Serrano-Pozo et al., 2021). Thus, the interactions we observed might reflect the accumulation of tau pathology with advancing age in APOE ε4 carriers. Interestingly, Van Etten et al. (2021) showed a negative association between age and hippocampal volume which was modulated by white matter hyper hyperintensities (WMH) in APOE ε4 carriers but not in non-carriers. These results provide an alternative explanation to our results, where vascular vulnerabilities would be the main driver of age-related atrophy in APOE ε4 carriers.

Our study has some limitations. First, the interactions we found between Aβ status and age as well as between APOE ε4 status and age on several MTL subregions were not strongly significant and did not survive multiple comparisons corrections. This might be explained by a lack of statistical power given the relatively small sample size. Thus, these analysis need to be replicated in a bigger cohort, potentially enriched in Aβ+ and APOE ε4 carriers individuals. However, it is important to keep in mind that the effects of amyloid and APOE on atrophy are certainly subtle, explaining the discrepancies in the literature. A strength of the present study is the use of high resolution T2-weighted images together with the ASHS algorithm. To our knowledge, this work is the first to investigate both the effects of amyloid and APOE on MTL subregions in the same sample using high resolution hippocampal images.

Altogether, our results provide new insight regarding age-related atrophy in these populations at-risk for AD. These results are important in the perspective of developing MRI-based biomarkers to detect early AD and further our understanding of MTL atrophy in normal and pathological ageing.

## Data Availability

All data produced in the present study are available upon reasonable request to the authors

## References

Adler DH, Wisse LEM, Ittyerah R, Pluta JB, Ding S-L, Xie L, Wang J, Kadivar S, Robinson JL, Schuck T, Trojanowski JQ, Grossman M, Detre JA, Elliott MA, Toledo JB, Liu W, Pickup S, Miller MI, Das SR, Wolk DA, Yushkevich PA. 2018. Characterizing the human hippocampus in aging and Alzheimer’s disease using a computational atlas derived from ex vivo MRI and histology. Proceedings of the National Academy of Sciences of the United States of America 115:4252–4257.

Baek MS, Cho H, Lee HS, Lee JH, Ryu YH, Lyoo CH. 2020. Effect of APOE ε4 genotype on amyloid-β and tau accumulation in Alzheimer’s disease. Alz Res Therapy 12:140.

Bourgeat P, Chételat G, Villemagne VL, Fripp J, Raniga P, Pike K, Acosta O, Szoeke C, Ourselin S, Ames D, Ellis KA, Martins RN, Masters CL, Rowe CC, Salvado O, AIBL Research Group. 2010. Beta-amyloid burden in the temporal neocortex is related to hippocampal atrophy in elderly subjects without dementia. Neurology 74:121–7.

Braak H, Braak E. 1991. Neuropathological stageing of Alzheimer-related changes. Acta neuropathologica 82:239–59.

Braak H, Braak E. 1995. Staging of alzheimer’s disease-related neurofibrillary changes. Neurobiology of Aging 16:271–278.

Burggren a C, Zeineh MM, Ekstrom a D, Braskie MN, Thompson PM, Small GW, Bookheimer SY. 2008. Reduced cortical thickness in hippocampal subregions among cognitively normal apolipoprotein E e4 carriers. NeuroImage 41:1177–83.

Cacciaglia R, Molinuevo JL, Falcón C, Brugulat-Serrat A, Sánchez-Benavides G, Gramunt N, Esteller M, Morán S, Minguillón C, Fauria K, Gispert JD, for the ALFA study. 2018. Effects of APOE-ε4 allele load on brain morphology in a cohort of middle-aged healthy individuals with enriched genetic risk for Alzheimer’s disease. Alzheimer’s & Dementia 14:902–912.

Chauveau L, Kuhn E, Palix C, Felisatti F, Ourry V, de La Sayette V, Chételat G, de Flores R. 2021. Medial Temporal Lobe Subregional Atrophy in Aging and Alzheimer’s Disease: A Longitudinal Study. Front Aging Neurosci 13:750154.

Delacourte A, David JP, Sergeant N, Buée L, Wattez A, Vermersch P, Ghozali F, Fallet-Bianco C, Pasquier F, Lebert F, Petit H, Di Menza C. 1999. The biochemical pathway of neurofibrillary degeneration in aging and Alzheimer’s disease. Neurology 52:1158–1165.

Dickerson BC, Bakkour A, Salat DH, Feczko E, Pacheco J, Greve DN, Grodstein F, Wright CI, Blacker D, Rosas HD, Sperling RA, Atri A, Growdon JH, Hyman BT, Morris JC, Fischl B, Buckner RL. 2009. The cortical signature of Alzheimer’s disease: regionally specific cortical thinning relates to symptom severity in very mild to mild AD dementia and is detectable in asymptomatic amyloid-positive individuals. Cerebral cortex (New York, NY : 1991) 19:497–510.

Dong Q, Zhang W, Wu J, Li B, Schron EH, McMahon T, Shi J, Gutman BA, Chen K, Baxter LC, Thompson PM, Reiman EM, Caselli RJ, Wang Y. 2019. Applying surface-based hippocampal morphometry to study APOE-E4 allele dose effects in cognitively unimpaired subjects. NeuroImage: Clinical 22:101744.

Donix M, Burggren AC, Suthana NA, Siddarth P, Ekstrom AD, Krupa AK, Jones M, Martin-Harris L, Ercoli LM, Miller KJ, Small GW, Bookheimer SY. 2010. Family history of Alzheimer’s disease and hippocampal structure in healthy people. The American journal of psychiatry 167:1399–406.

Dounavi M-E, Mak E, Wells K, Ritchie K, Ritchie CW, Su L, O’ Brien JT. 2020. Volumetric alterations in the hippocampal subfields of subjects at increased risk of dementia. Neurobiology of Aging 91:36–44.

Eichenbaum H. 2017. The role of the hippocampus in navigation is memory. Journal of Neurophysiology 117:1785–1796.

de Flores R, La Joie R, Chételat G. 2015a. Structural imaging of hippocampal subfields in healthy aging and Alzheimer’s disease. Neuroscience 309:29–50.

de Flores R, La Joie R, Landeau B, Perrotin A, Mézenge F, de La Sayette V, Eustache F, Desgranges B, Chételat G. 2015b. Effects of age and Alzheimer’s disease on hippocampal subfields: comparison between manual and FreeSurfer volumetry. Human brain mapping 36:463–74.

Fouquet M, Besson FL, Gonneaud J, La Joie R, Chételat G. 2014. Imaging brain effects of APOE4 in cognitively normal individuals across the lifespan. Neuropsychology review 24:290–9.

Gertje EC, Pluta J, Das S, Mancuso L, Kliot D, Yushkevich P, Wolk D. 2016. Clinical Application of Automatic Segmentation of Medial Temporal Lobe Subregions in Prodromal and Dementia-Level Alzheimer’s Disease. Journal of Alzheimer’s disease : JAD:1–11.

Geuze E, Vermetten E, Bremner JD. 2005. MR-based in vivo hippocampal volumetrics: 2. Findings in neuropsychiatric disorders. Molecular psychiatry 10:160–84.

Hata K, Nakamoto K, Nunomura A, Sone D, Maikusa N, Ogawa M, Sato N, Matsuda H. 2019. Automated Volumetry of Medial Temporal Lobe Subregions in Mild Cognitive Impairment and Alzheimer Disease. Alzheimer Disease & Associated Disorders 33:206– 211.

Hodgetts CJ, Voets NL, Thomas AG, Clare S, Lawrence AD, Graham KS. 2017. Ultra-High-Field fMRI Reveals a Role for the Subiculum in Scene Perceptual Discrimination. The Journal of neuroscience : the official journal of the Society for Neuroscience 37:3150– 3159.

Honea RA, Vidoni E, Harsha A, Burns JM. 2009. Impact of APOE on the healthy aging brain: a voxel-based MRI and DTI study. J Alzheimers Dis 18:553–564.

Hsu PJ, Shou H, Benzinger T, Marcus D, Durbin T, Morris JC, Sheline YI. 2015. Amyloid burden in cognitively normal elderly is associated with preferential hippocampal subfield volume loss. Journal of Alzheimer’s disease : JAD 45:27–33.

Jack CR, Bennett DA, Blennow K, Carrillo MC, Dunn B, Haeberlein SB, Holtzman DM, Jagust W, Jessen F, Karlawish J, Liu E, Molinuevo JL, Montine T, Phelps C, Rankin KP, Rowe CC, Scheltens P, Siemers E, Snyder HM, Sperling R, Elliott C, Masliah E, Ryan L, Silverberg N. 2018. NIA-AA Research Framework: Toward a biological definition of Alzheimer’s disease. Alzheimer’s & Dementia 14:535–562.

Jagust W. 2018. Imaging the evolution and pathophysiology of Alzheimer disease. Nature Reviews Neuroscience 19:687–700.

Kerchner GA, Berdnik D, Shen JC, Bernstein JD, Fenesy MC, Deutsch GK, Wyss-Coray T, Rutt BK. 2014. APOE ε4 worsens hippocampal CA1 apical neuropil atrophy and episodic memory. Neurology 82:691–7.

Khan W, Giampietro V, Banaschewski T, Barker GJ, Bokde ALW, Büchel C, Conrod P, Flor H, Frouin V, Garavan H, Gowland P, Heinz A, Ittermann B, Lemaître H, Nees F, Paus T, Pausova Z, Rietschel M, Smolka MN, Ströhle A, Gallinat J, Vellas B, Soininen H, Kloszewska I, Tsolaki M, Mecocci P, Spenger C, Villemagne VL, Masters CL, Muehlboeck J-S, Bäckman L, Fratiglioni L, Kalpouzos G, Wahlund L-O, Schumann G, Lovestone S, Williams SCR, Westman E, Simmons A, Alzheimer–s Disease Neuroimaging Initiative, AddNeuroMed Consortium, Australian, Imaging, Biomarkers, and Lifestyle Study Research Group, IMAGEN consortium. 2017. A Multi-Cohort Study of ApoE ε4 and Amyloid-β Effects on the Hippocampus in Alzheimer’s Disease. J Alzheimers Dis 56:1159–1174.

La Joie R, Visani AV, Lesman-Segev OH, Baker SL, Edwards L, Iaccarino L, Soleimani-Meigooni DN, Mellinger T, Janabi M, Miller ZA, Perry DC, Pham J, Strom A, Gorno-Tempini ML, Rosen HJ, Miller BL, Jagust WJ, Rabinovici GD. 2021. Association of APOE4 and Clinical Variability in Alzheimer Disease With the Pattern of Tau- and Amyloid-PET. Neurology 96:e650–e661.

Lace G, Savva GM, Forster G, De Silva R, Brayne C, Matthews FE, Barclay JJ, Dakin L, Ince PG, Wharton SB. 2009. Hippocampal tau pathology is related to neuroanatomical connections: An ageing population-based study. Brain 132:1324–1334.

Lind J, Larsson A, Persson J, Ingvar M, Nilsson L-G, Bäckman L, Adolfsson R, Cruts M, Sleegers K, Van Broeckhoven C, Nyberg L. 2006. Reduced hippocampal volume in non-demented carriers of the apolipoprotein E ε4: Relation to chronological age and recognition memory. Neuroscience Letters 396:23–27.

Maass A, Landau S, Baker SL, Horng A, Lockhart SN, La Joie R, Rabinovici GD, Jagust WJ. 2017. Comparison of multiple tau-PET measures as biomarkers in aging and Alzheimer’s disease. NeuroImage 157:448–463.

Martí-Juan G, Sanroma-Guell G, Cacciaglia R, Falcon C, Operto G, Molinuevo JL, González Ballester MÁ, Gispert JD, Piella G, Initiative TADN, Study TA. 2021. Nonlinear interaction between APOE ε4 allele load and age in the hippocampal surface of cognitively intact individuals. Human Brain Mapping 42:47–64.

Mattsson N, Insel PS, Nosheny R, Tosun D, Trojanowski JQ, Shaw LM, Jack CR, Donohue MC, Weiner MW. 2014. Emerging β-Amyloid Pathology and Accelerated Cortical Atrophy. JAMA Neurol 71:725.

Moscovitch M, Nadel L, Winocur G, Gilboa A, Rosenbaum RS. 2006. The cognitive neuroscience of remote episodic, semantic and spatial memory. Current Opinion in Neurobiology 16:179–190.

Mueller SG, Schuff N, Raptentsetsang S, Elman J, Weiner MW. 2008. Selective effect of Apo e4 on CA3 and dentate in normal aging and Alzheimer’s disease using high resolution MRI at 4 T. NeuroImage 42:42–8.

Mueller SG, Yushkevich PA, Das S, Wang L, Van Leemput K, Iglesias JE, Alpert K, Mezher A, Ng P, Paz K, Weiner MW. 2018. Systematic comparison of different techniques to measure hippocampal subfield volumes in ADNI2. NeuroImage: Clinical 17:1006–1018.

Nosheny RL, Insel PS, Mattsson N, Tosun D, Buckley S, Truran D, Schuff N, Aisen PS, Weiner MW. 2019. Associations among amyloid status, age, and longitudinal regional brain atrophy in cognitively unimpaired older adults. Neurobiology of Aging 82:110–119.

Oh H, Madison C, Villeneuve S, Markley C, Jagust WJ. 2014. Association of Gray Matter Atrophy with Age, β-Amyloid, and Cognition in Aging. Cerebral Cortex 24:1609–1618.

Parker TD, Cash DM, Lane CAS, Lu K, Malone IB, Nicholas JM, James S-N, Keshavan A, Murray-Smith H, Wong A, Buchanan SM, Keuss SE, Sudre CH, Modat M, Thomas DL, Crutch SJ, Richards M, Fox NC, Schott JM. 2019. Hippocampal subfield volumes and pre-clinical Alzheimer’s disease in 408 cognitively normal adults born in 1946. PLOS ONE 14:e0224030.

Pereira JB, Harrison TM, La Joie R, Baker SL, Jagust WJ. 2020. Spatial patterns of tau deposition are associated with amyloid, ApoE, sex, and cognitive decline in older adults. Eur J Nucl Med Mol Imaging 47:2155–2164.

Pini L, Pievani M, Bocchetta M, Altomare D, Bosco P, Cavedo E, Galluzzi S, Marizzoni M, Frisoni GB. 2016. Brain atrophy in Alzheimer’s Disease and aging. Ageing research reviews 30:1–24.

Poisnel G, Arenaza-Urquijo E, Collette F, Klimecki OM, Marchant NL, Wirth M, de La Sayette V, Rauchs G, Salmon E, Vuilleumier P, Frison E, Maillard A, Vivien D, Lutz A, Chételat G. 2018. The Age-Well randomized controlled trial of the Medit-Ageing European project: Effect of meditation or foreign language training on brain and mental health in older adults. Alzheimer’s and Dementia: Translational Research and Clinical Interventions 4:714–723.

Ramanan VK, Castillo AM, Knopman DS, Graff-Radford J, Lowe VJ, Petersen RC, Jack CR, Mielke MM, Vemuri P. 2019. Association of Apolipoprotein E ε4, Educational Level, and Sex With Tau Deposition and Tau-Mediated Metabolic Dysfunction in Older Adults. JAMA Netw Open 2:e1913909.

Reiman EM, Uecker A, Caselli RJ, Lewis S, Bandy D, De Leon MJ, De Santi S, Convit A, Osborne D, Weaver A, Thibodeau SN. 1998. Hippocampal volumes in cognitively normal persons at genetic risk for Alzheimer’s disease. Ann Neurol 44:288–291.

Reiter K, Nielson KA, Durgerian S, Woodard JL, Smith JC, Seidenberg M, Kelly DA, Rao SM. 2017. Five-Year Longitudinal Brain Volume Change in Healthy Elders at Genetic Risk for Alzheimer’s Disease. J Alzheimers Dis 55:1363–1377.

Salvadó G, Grothe MJ, Groot C, Moscoso A, Schöll M, Gispert JD, Ossenkoppele R, for the Alzheimer’s Disease Neuroimaging Initiative. 2021. Differential associations of APOE-ε2 and APOE-ε4 alleles with PET-measured amyloid-β and tau deposition in older individuals without dementia. Eur J Nucl Med Mol Imaging 48:2212–2224.

Serrano-Pozo A, Das S, Hyman BT. 2021. APOE and Alzheimer’s disease: advances in genetics, pathophysiology, and therapeutic approaches. The Lancet Neurology 20:68–80.

Small S a, Schobel S a, Buxton RB, Witter MP, Barnes C a. 2011. A pathophysiological framework of hippocampal dysfunction in ageing and disease. Nature reviews Neuroscience 12:585–601.

Squire LR, Stark CEL, Clark RE. 2004. the Medial Temporal Lobe. Annual Review of Neuroscience 27:279–306.

Storandt M, Mintun MA, Head D, Morris JC. 2009. Cognitive Decline and Brain Volume Loss as Signatures of Cerebral Amyloid-β Peptide Deposition Identified With Pittsburgh Compound B: Cognitive Decline Associated With Aβ Deposition. Arch Neurol [Internet] 66. Available from: http://archneur.jamanetwork.com/article.aspx?doi=10.1001/archneurol.2009.272

Therriault J, Benedet AL, Pascoal TA, Mathotaarachchi S, Chamoun M, Savard M, Thomas E, Kang MS, Lussier F, Tissot C, Parsons M, Qureshi MNI, Vitali P, Massarweh G, Soucy J-P, Rej S, Saha-Chaudhuri P, Gauthier S, Rosa-Neto P. 2020. Association of Apolipoprotein E ε4 With Medial Temporal Tau Independent of Amyloid-β. JAMA Neurol 77:470.

Tohgi H, Takahashi S, Kato E, Homma A, Niina R, Sasaki K, Yonezawa H, Sasaki M. 1997. Reduced size of right hippocampus in 39-to 80-year-old normal subjects carrying the apolipoprotein E epsilon4 allele. Neurosci Lett 236:21–24.

Trivedi MA, Schmitz TW, Ries ML, Torgerson BM, Sager MA, Hermann BP, Asthana S, Johnson SC. 2006. Reduced hippocampal activation during episodic encoding in middle-aged individuals at genetic risk of Alzheimer’s Disease: a cross-sectional study. BMC Med 4:1.

Tzioras M, Davies C, Newman A, Jackson R, Spires-Jones T. 2019. Invited Review: APOE at the interface of inflammation, neurodegeneration and pathological protein spread in Alzheimer’s disease. Neuropathol Appl Neurobiol 45:327–346.

Van Etten EJ, Bharadwaj PK, Hishaw GA, Huentelman MJ, Trouard TP, Grilli MD, Alexander GE. 2021. Influence of regional white matter hyperintensity volume and apolipoprotein E ε4 status on hippocampal volume in healthy older adults. Hippocampus 31:469–480.

Veldsman M, Nobis L, Alfaro-Almagro F, Manohar S, Husain M. 2021. The human hippocampus and its subfield volumes across age, sex and APOE e4 status. Brain Communications 3:fcaa219.

Vilor-Tejedor N, Evans TE, Adams HH, González-de-Echávarri JM, Molinuevo JL, Guigo R, Gispert JD, Operto G. 2021. Genetic Influences on Hippocampal Subfields: An Emerging Area of Neuroscience Research. Neurol Genet 7:e591.

Voineskos AN, Winterburn JL, Felsky D, Pipitone J, Rajji TK, Mulsant BH, Chakravarty MM. 2015. Hippocampal (subfield) volume and shape in relation to cognitive performance across the adult lifespan. Human brain mapping 36:3020–37.

Whitwell JL, Tosakulwong N, Weigand SD, Senjem ML, Lowe VJ, Gunter JL, Boeve BF, Knopman DS, Dickerson BC, Petersen RC, Jack CR. 2013. Does amyloid deposition produce a specific atrophic signature in cognitively normal subjects? NeuroImage: Clinical 2:249–257.

Wishart HA, Saykin AJ, McAllister TW, Rabin LA, McDonald BC, Flashman LA, Roth RM, Mamourian AC, Tsongalis GJ, Rhodes CH. 2006. Regional brain atrophy in cognitively intact adults with a single APOE 4 allele. Neurology 67:1221–1224.

Wisse LE, Xie L, Das SR, de Flores R, Hansson O, Habes M, Doshi J, Davatzikos C, Yushkevich PA, Wolk DA. 2022. Tau pathology mediates age effects on medial temporal lobe structure. Neurobiology of Aging 109:135–144.

Wisse LEM, Chételat G, Daugherty AM, Flores R, Joie R, Mueller SG, Stark CEL, Wang L, Yushkevich PA, Berron D, Raz N, Bakker A, Olsen RK, Carr VA. 2021. Hippocampal subfield volumetry from structural isotropic 1 mm3 MRI scans: A note of caution. Hum Brain Mapp 42:539–550.

Wolk DA, Das SR, Mueller SG, Weiner MW, Yushkevich PA. 2017. Medial temporal lobe subregional morphometry using high resolution MRI in Alzheimer’s disease. Neurobiology of Aging 49:204–213.

Xie L, Wisse LEM, Pluta J, de Flores R, Piskin V, Manjón JV, Wang H, Das SR, Ding S-L, Wolk DA, Yushkevich PA. 2019. Automated segmentation of medial temporal lobe subregions on in vivo T1-weighted MRI in early stages of Alzheimer’s disease. Human Brain Mapping 40:3431–3451.

Yushkevich PA, Pluta JB, Wang H, Xie L, Ding S-L, Gertje EC, Mancuso L, Kliot D, Das SR, Wolk D a. 2015. Automated volumetry and regional thickness analysis of hippocampal subfields and medial temporal cortical structures in mild cognitive impairment. Human brain mapping 36:258–87.

