## Supplementary Table 1 for "Effects of amyloid and *APOE4* on medial temporal lobe subregions in cognitively unimpaired elderly"

**Supplementary Table 1 : Effects of amyloid and APOE4 status on MTL subregions**

| **a) Amyloid Status** |  |  |  |  |  |  |
| --- | --- | --- | --- | --- | --- | --- |
|  |  |  | Group comparison (Aβ- vs Aβ+) | | Interaction (Aβ status * age) | |
|  | Aβ- Volume | Aβ+ Volume | F | P | F | P |
| CA1 | 0.96 (0.10) | 0.93 (0.14) | 0.22 | 0.64 | **4.16** | **0.04** |
| CA2 | 0.02 (0.003) | 0.02 (0.004) | 2.51 | 0.12 | 0.44 | 0.51 |
| CA3 | 0.05 (0.01) | 0.06 (0.01) | **6.12** | **0.01** | 0.15 | 0.69 |
| DG | 0.42 (0.05) | 0.4 (0.07) | 0.40 | 0.53 | 1.90 | 0.17 |
| SUB | 0.32 (0.04) | 0.32 (0.04) | 0.43 | 0.51 | 1.17 | 0.28 |
| ERC | 0.41 (0.04) | 0.4 (0.07) | 0.53 | 0.47 | *3.54* | *0.06* |
| BA35 | 0.36 (0.06) | 0.35 (0.04) | 0.27 | 0.60 | 1.63 | 0.20 |
| BA36 | 1.24 (0.18) | 1.25 (0.19) | 0.36 | 0.55 | 1.47 | 0.23 |
| PHC | 0.67 (0.09) | 0.66 (0.09) | 0.02 | 0.87 | *3.62* | *0.06* |
| Whole hippocampus | 1.77 (0.18) | 1.73 (0.24) | 0.19 | 0.67 | *3.30* | *0.07* |
| **b) APOE Status** |  |  |  |  |  |  |
|  |  |  | Group comparison (ε4- vs ε4+) | | Interaction (APOE status * age) | |
|  | ε4- Volume | ε4+ Volume | F | P | F | P |
| CA1 | 0.96 (0.10) | 0.93 (0.13) | 0.69 | 0.41 | **4.07** | **0.04** |
| CA2 | 0.02 (0.003) | 0.02 (0.004) | 1.20 | 0.27 | 0.14 | 0.71 |
| CA3 | 0.05 (0.01) | 0.05 (0.01) | 2.90 | 0.09 | 0.08 | 0.77 |
| DG | 0.41 (0.05) | 0.41 (0.07) | 0.04 | 0.84 | *3.45* | *0.07* |
| SUB | 0.32 (0.04) | 0.32 (0.04) | 0.02 | 0.89 | **4.81** | **0.03** |
| ERC | 0.41 (0.04) | 0.4 (0.06) | 0.01 | 0.96 | **4.13** | **0.04** |
| BA35 | 0.37 (0.06) | 0.34 (0.06) | 2.83 | 0.09 | 0.36 | 0.55 |
| BA36 | 1.24 (0.16) | 1.26 (0.22) | 8,00E-04 | 0.99 | *3.08* | *0.08* |
| PHC | 0.67 (0.08) | 0.66 (0.1) | 0.03 | 0.86 | 2.63 | 0.11 |
| Whole hippocampus | 1.77 (0.17) | 1.73 (0.24) | 0.47 | 0.49 | **4.75** | **0.03** |
